# Precision MRI Phenotyping Enables Detection of Small Changes in Body Composition for Longitudinal Cohorts

**DOI:** 10.1101/2021.08.29.21262790

**Authors:** Brandon Whitcher, Marjola Thanaj, Madeleine Cule, Yi Liu, Nicolas Basty, Elena P. Sorokin, Jimmy D. Bell, E. Louise Thomas

## Abstract

Longitudinal studies provide unique insights into the impact of environmental factors and lifespan issues on health and disease. Here we investigate changes in body composition in 3,088 free-living participants, part of the UK Biobank in-depth imaging study. All participants underwent neck-to-knee MRI scans at the first imaging visit and after approximately two years (second imaging visit). Image-derived phenotypes for each participant were extracted using a fully-automated image processing pipeline, including volumes of several tissues and organs: liver, pancreas, spleen, kidneys, total skeletal muscle, iliopsoas muscle, visceral adipose tissue (VAT), abdominal subcutaneous adipose tissue (ASAT), as well as fat and iron content in liver, pancreas and spleen. Overall, no significant changes were observed in BMI, body weight, or waist circumference over the scanning interval, despite some large individual changes. A significant decrease in grip strength was observed, coupled to small, but statistically significant, decrease in all skeletal muscle measurements. Significant increases in VAT and intermuscular fat in the thighs were also detected in the absence of changes in BMI, waist circumference and ectopic-fat deposition. Adjusting for disease status at the first imaging visit did not have an additional impact on the changes observed. In summary, we show that even after a relatively short period of time significant changes in body composition can take place, probably reflecting the obesogenic environment currently inhabited by most of the general population in the United Kingdom.

## Introduction

The contribution of longitudinal cohort studies to our understanding of the development of disease and the impact of lifestyle cannot be underestimated. Forward thinking studies such as the Framingham Heart Study have revolutionised our understanding of the development and epidemiology of cardiovascular disease^1^. Investigations aimed at understanding the relationship between body composition and lifestyle at a population scale have typically been explored in cross-sectional studies, or using relatively simple/cost-effective measurements such as body mass index (BMI), anthropometry and questionnaires^2^. However, precise phenotypic measurements of detailed body composition including multiple organ volumes, subdivisions of adipose tissue and the composition and quality of tissues, has been limited to relatively small studies or few measurements. Long-term cohort studies tracking changes in adipose tissue and ectopic fat such as KORA (Cooperative Health Research in the Region of Augsburg) or Study of Health in Pomerania (SHIP), which includes whole-body MRI, have only obtained MRI measurements at a single time point^3,4^. Others such as the Dallas Heart Study^5^ undertook longitudinal MRI that included whole-body and liver fat measurements, but like other cohorts this study focussed on heart and vessel health and did not make detailed measurements of other abdominal organs.

The advent of deep learning methods, particularly applied to the analysis of large MRI datasets, has made the measurement of multiple abdominal organs at scale a possibility. This is essential in light of the growing number of population-based cohorts world wide such as the UK Biobank^6^ and German National Cohort^7^. The UK Biobank imaging study, originally designed to be cross-sectional with 100,000 participants has now expanded its original remit to include an additional imaging visit for the whole cohort, approximately four to five years after the initial scan^8^. In addition a subset of 10,000 UK Biobank participants are having a follow-up scan approximately two years after their first imaging visit (Dementias Platform UK). The longitudinal assessment of changes in organ health in a large cohort is now possible.

To date 3,209 participants have undergone MR imaging at the first imaging visit and again after approximately a two-year period. The primary aim of our study was to determine whether there would be detectable longitudinal changes in organ volume and composition in a large free-living population living in an obesogenic environment. Our secondary aim was to determine whether these changes would be accelerated or ameliorated in individuals living with a chronic disease or physiological condition. For the purpose of this study conditions were selected based on incidence within the UK Biobank population and/or known impact on organs of interest. Here we describe our initial observations of these data. We generated a total of 27 image derived phenotypes (IDPs) from the MR abdominal protocol and combined these with physiological measures with reference to disease/physiological condition. We undertook exploratory data analysis prior to identifying combinations of IDPs and disease for statistical modelling. We found that the trajectory of longitudinal changes in body-composition IDPs was dependant on the physiological state of the participants.

## Methods

### Data

Approximately 49,000 participants have received their first MRI scans (brain, heart and abdominal area) as of December 15, 2020, with 3,209 having undergone a follow-up imaging visit. UK Biobank participants undergoing repeat scanning were part of a cohort of 10,000 participants recruited by the Dementia Platform UK study to include 5,000 selected based on dementia risk factors which are known to the participants, namely increasing age and family history of dementia (self-reported by participants) and 5,000 participants without family history of dementia. Participant age at the second imaging visit was required to be 55 years or older (range 55 – 85 years), with the aim of achieving an eventual mean cohort age of approximately 70 years. For the current study of body composition measurements, we were blinded as to whether people had a family history of dementia. Participants with imaging data from the abdominal protocol at both time points have been included in this manuscript; only 3,088 met this requirement. Weight using a Tanita BC418MA body composition analyser, standing height using a Seca 240 Height Measure, waist (narrowest part of the trunk) and hip (widest part of the trunk) circumference using a Seca 200 tape measure, grip strength using a Jamar J00105 hydraulic hand dynamometer and seated blood pressure using an Omron 705 IT electronic blood pressure monitor were all measured as a part of the UK Biobank assessment at each visit.

Participant data from the UKBB cohort was obtained as previously described^6^ through UKBB Access Application number 44584. The UKBB has approval from the North West Multi-Centre Research Ethics Committee (REC reference: 11/NW/0382). All methods were performed in accordance with the relevant guidelines and regulations, and informed consent was obtained from all participants. Researchers may apply to use the UKBB data resource by submitting a health-related research proposal that is in the public interest. More information may be found on the UKBB researchers and resource catalogue pages (www.ukbiobank.ac.uk).

### Disease/physiological condition categories

Disease categories of interest including cardiovascular, liver and kidney, type-1 and type-2 diabetes, metabolic and cancer were selected given the extensive evidence that changes in body composition and organ health have been associated with them, as well as their frequency within the UK Biobank. Menopause, while not a disease, was selected as a physiological state, likely to influence body composition. Disease categories were defined based on recorded hospital episode statistics (HES) data and self-reported information. If the International Classification of Diseases 9th and 10th edition (ICD9 and ICD10, respectively) for HES data, or self-reported disease codes (UK Biobank fields) were reported at least once for each participant, at the time and before the first imaging visit, they were classified as a case. A summary of categories and the number of participants diagnosed with each disease/physiological condition, is shown in Table 1.

**Table 1.** Number of participants diagnosed by disease or physiological condition on and before their first imaging visit out of the 3,088 included in the study.

| Disease or Physiological Condition | N |
| --- | --- |
| Cardiovascular Disease | 214 |
| Liver Disease | 38 |
| Kidney Disease | 18 |
| Type-1 Diabetes | 14 |
| Type-2 Diabetes | 147 |
| Metabolic Disorder | 199 |
| Cancer | 451 |
| Menopause | 1,351 |

The codes corresponding to the considered disease/physiological condition traits are provided in Supplementary Table S2. If a cell is empty in the table, it means that no code for that category has been used. The ICD codes for cardiovascular disease (CVD) were defined from Lees *et al*.^9^ and Tavaglione *et al*.^10^, and self-reported codes used were “angina”,”heart attack/myocardial infraction”, “stroke”, “subarachnoid haemorrhage”, “brain haemorrhage”, “ischaemic stroke” and “heart failure/pulmonary odema”. The ICD codes for liver disease were taken from Schneider *et al*.^11^ and the self-reported codes used were “liver failure/cirrhosis”, “infective/viral hepatitis”, “alcoholic liver disease/alcoholic cirrhosis” and “liver/biliary/pancreas problem”. The codes for kidney disease were taken from Lin *et al*.^12^ and *Definitions of End Stage Renal Disease for UK Biobank Phase 1 Outcomes Adjudication* (biobank.ndph.ox.ac.uk/ukb/ukb/docs/alg_outcome_esrd.pdf). The codes for type-1 diabetes were selected based on the ICD codes for “insulin-dependent diabetes mellitus” and the self-reported code for “type-1 diabetes”. The codes for type-2 diabetes (T2DM) were selected based on the ICD codes for “non-insulin-dependent diabetes mellitus” and the self-reported code for “diabetes” and “type-2 diabetes”. Cancer was defined from all the types of cancer in ICD9 and ICD10 and their self-reported fields 20001 and 2453. The codes for metabolic disorder corresponding to carbohydrates and lipids were taken from the ICD category “metabolic disorders”. Menopause was defined based on the ICD category “menopausal and female climacteric states” and their self-reported fields 20002 and 2724.

### Image acquisition and analysis

Full details regarding the UK Biobank MR abdominal protocol have previously been reported^8^. The data included in this paper focused on the neck-to-knee Dixon MRI acquisition and separate single-slice multiecho MRI acquisitions for the liver and pancreas. All data were analyzed using our dedicated image processing pipelines for the Dixon and single-slice multiecho acquisitions. Deep learning algorithms were used to segment organs and tissue^13,14^. Proton density fat fraction (PDFF) and R2* were calculated from the Phase Regularized Estimation using Smoothing and Constrained Optimization (PRESCO) method^15^. Conversion from R2* to iron concentration followed McKay *et al*.^16^.

The following organs and tissue were segmented in 3D and summarized by volume: liver, lungs, pancreas, left kidney, right kidney, spleen, ASAT, VAT, subcutaneous adipose tissue in the thighs, internal fat in the thighs, total skeletal muscle, total iliopsoas muscle, total thigh muscle and heart. The following organs and tissue were segmented in 2D and summarized by cross-sectional area (CSA): total paraspinal muscle, superior vena cava, aortic arch, descending thoracic aorta, descending suprarenal aorta and descending infrarenal aorta. Median PDFF was calculated for the following organs and tissue in 2D: liver, pancreas and paraspinal muscle. Median iron concentration was calculated for the following organs and tissue in 2D: liver, pancreas, paraspinal muscle and spleen. Where left and right locations are specified, separate IDPs are provided for the left and right. If the word “total” was used, then the left and right locations have been combined into a single IDP. Total skeletal muscle refers to all skeletal muscle tissue in the neck-to-knee anatomical coverage of the Dixon MRI acquisition.

Vascular CSAs were obtained by applying a cut plane to the 3D segmentations of the aorta and vena cava. As vessel volume on its own is not very informative, because the beginning and ending locations are not well defined, we characterised vessels via CSA at several clinically-relevant landmarks^17^. We placed the landmarks based on anatomical definitions using organ segmentations. The aortic arch was placed at the top of the aorta segmentation, the thoracic aorta at the centre of mass of the heart segmentation, the suprarenal aorta at the top of the kidney segmentation, the infrarenal aorta at the bottom of the kidney segmentation and the superior vena cava at the top of the heart segmentation. We used those landmarks to compute tangents and derive orthogonal planes to account for the fact that vessels do not simply follow the body in a vertical and perpendicular fashion^18^. CSA was calculated for each section of the vessel based on the intersection of the 3D vessel segmentation and the orthogonal plane.

Quality control procedures were applied to all IDPs by assessing the univariate distributions and bivariate distribution of the two visits. Visual inspection of the MRI data was performed to determine the cause of extreme values in the IDPs and confirm exclusion. Three participants were excluded due to a high level of noise throughout the acquisition at second imaging visit. The left kidney volume was excluded for three participants and the right kidney volume was excluded for three different participants, with a total of six kidney-volume IDPs excluded. One heart volume and one lung volume were excluded, from different participants. The spleen volume was excluded from five participants. One VAT volume and one liver PDFF measurement were excluded, from different participants. Minimum and maximum thresholds for the CSA of the aortic arch and superior vena cava were set at 150 mm^2^ and 1, 600 mm^2^, respectively. A total of 33 aortic arch volumes and 66 superior vena cava volumes were excluded because of these thresholds, all from different participants. The pancreas volume was excluded from 36 subjects, along with the corresponding PDFF and iron concentration IDPs, due to a failed segmentation procedure in at least one of the imaging time points.

### Statistical analysis

All summary statistics, hypothesis tests, regression models and figures were performed using the R software environment for statistical computing and graphics^19^. Descriptive statistics are expressed as mean and standard deviation in all tables and in the text. Variables were tested for normality using the Shapiro-Wilk’s test, the null hypothesis was rejected in all cases. Spearman’s rank correlation coefficient was used to assess monotonic trends between variables. The Wilcoxon rank-sum test was used to compare means between groups, and the Wilcoxon signed-rank test with paired observations. The threshold for statistical significance of *p*-values was adjusted for the number of formal hypothesis tests performed in Table 3, Table 5, Supplementary Table S1 and Supplementary Table S3. The Bonferroni-corrected threshold was 0.05*/*179 = 2.8×10^*−*4^.

We computed correlation matrices between the IDPs and clinical/demographic variables under the disease and physiological condition categories identified in Table 1. Relationships between variables were assessed and seven combinations were identified for further study (Table 2). Linear mixed-effects models, with these specific IDP and disease category combinations, were fit to the data using the **lme4** package^20^, where the IDP was the response and the disease category a fixed effect. Data from both time points were included in the linear mixed-effects model instead of performing an analysis on the differences^21^. Participant IDs were included as a random effect (intercept only) given that repeated measurements were obtained. Additional fixed effects included in all models were: age at the first imaging visit, gender, BMI, assessment center, systolic blood pressure, diastolic blood pressure, volume of ASAT and volume of VAT. We also included fixed effects depending on which IDP was in the model. Specifically, liver PDFF and iron concentration were added to the liver volume model and grip strength of the non-dominant hand was added to the total-muscle and iliopsoas-muscle volume models. All continuous variables used as fixed effects were centered to aid in the interpretation of the estimated regression coefficients. To investigate the effect of time on participants in different disease categories, an indicator variable for the visit was crossed with disease category so that both the main effects of visit and disease/condition were included in addition to their interaction term. P-values for the regression coefficients in the linear mixed-effects models were computed using the Satterthwaite approximation to the degrees of freedom for the *t*-statistic^22^ as part of the **lmerTest** package^23^. The intraclass correlation coefficient (ICC), adjusted for fixed effects in the linear mixed-effects model, was calculated as a measure of repeatability^24^ for the IDPs using the **performance** package^25^.

**Table 2.** Linear mixed-effects models by number, specifying the image derived phenotype (IDP) and disease category.

| Model | Image Derived Phenotype | Disease |
| --- | --- | --- |
| 1 | Total Muscle Volume | Type-2 Diabetes |
| 2 | Total Iliopsoas Muscle Volume | Type-2 Diabetes |
| 3 | Total Muscle Volume | Cardiovascular Disease |
| 4 | Total Kidney Volume | Cardiovascular Disease |
| 5 | Liver Volume | Metabolic Disorder |
| 6 | Total Lung Volume | Liver Disease |
| 7 | Total Kidney Volume | Liver Disease |

Diagnostics were performed to assess the linear mixed-effects models. Plotting the residuals versus fitted values for the continuous variables did not uncover any deviations from a linear form. Constant variance was also observed across the fitted range. There were deviations from normality in the residuals at both tails of the distribution for all models. This is not surprising since the IDPs must be non-negative and, hence, are slightly skewed to the right.

## Results

Of the 3,209 participants who underwent second scans, 3,088 had complete imaging data at both visits from the abdominal protocol. The demographics of this cohort are described in Table 3. Of these participants, 1, 838 (59.5%) were scanned at the Cheadle and 1, 250 (40.5%) at the Newcastle UK Biobank sites. The female-to-male ratio was 50.1:49.9, the average age of male participants was 63.49 ± 7.28 years at the first imaging visit and for female participants it was 62.35 ± 7.15 years. The average BMI of the male participants was 26.66 ± 3.82 kg/m^2^ (range 18.32−46.14 kg/m^2^) at the first imaging visit and for female participants 25.82 ± 4.40 kg/m^2^ (range 13.39−50.61 kg/m^2^), with the average value in both groups being categorized as slightly overweight. There were small differences between the characteristics at the first imaging visit of the longitudinal subset and the wider UK Biobank imaging cohort (Supplementary Table S1), which may be accounted for by the inclusion criteria for this specific cohort (see Methods).

**Table 3.** Demographics for participants in the longitudinal cohort, separated by gender. Values are reported as mean and standard deviation. An asterisk (*) indicates the *p*-value is below the significance threshold adjusted for multiple comparisons. BMI: body mass index; BP: blood pressure.

|  | Women |  |  |  |  | *p*-value |
| --- | --- | --- | --- | --- | --- | --- |
|  | First Imaging Visit |  | Second Imaging Visit |  | Difference |  |
| *N* | 1,548 |  |  |  |  |  |
| *N* (Cheadle) | 888 |  |  |  |  |  |
| *N* (Newcastle) | 660 |  |  |  |  |  |
| Age | 1,548 | 62.35 ± 7.15 | 1,548 | 64.62 ± 7.15 | 2.27 ± 0.44 |  |
| Weight (kg) | 1,548 | 68.44 ± 12.09 | 1,548 | 68.35 ± 12.29 | −0.09 ± 3.57 | 0.8544 |
| Height (cm) | 1,548 | 162.81 ± 6.20 |  |  |  |  |
| BMI (kg/m<sup>2</sup>) | 1,548 | 25.82 ± 4.40 | 1,548 | 25.79 ± 4.48 | −0.03 ± 1.34 | 0.878 |
| Waist Circumference (cm) | 1,471 | 82.66 ± 11.42 | 1,543 | 82.99 ± 11.24 | 0.29 ± 6.59 | 0.074 |
| Hip Circumference (cm) | 1,471 | 100.78 ± 9.36 | 1,543 | 99.74 ± 8.94 | −0.99 ± 5.51 | 2.1 × 10<sup>−11</sup>* |
| Waist-to-Hip Ratio | 1,471 | 0.82 ± 0.07 | 1,543 | 0.83 ± 0.07 | 0.01 ± 0.06 | 5.3 × 10<sup>−11</sup>* |
| Systolic BP (mm Hg) | 1,071 | 135.38 ± 18.29 | 1,415 | 138.98 ± 19.35 | 3.79 ± 14.88 | 1 × 10<sup>−14</sup>* |
| Diastolic BP (mm Hg) | 1,071 | 75.87 ± 9.68 | 1,415 | 77.70 ± 9.60 | 1.76 ± 7.95 | 6.7 × 10<sup>−12</sup>* |
| Grip Strength (kg) | 1,452 | 21.79 ± 5.66 | 1,520 | 20.77 ± 5.97 | −1.15 ± 5.55 | 2 × 10<sup>−14</sup>* |
|  | Men |  |  |  |  | *p*-value |
|  | First Imaging Visit |  | Second Imaging Visit |  | Difference |  |
| *N* | 1,540 |  |  |  |  |  |
| *N* (Cheadle) | 950 |  |  |  |  |  |
| *N* (Newcastle) | 590 |  |  |  |  |  |
| Age | 1,540 | 63.49 ± 7.28 | 1,540 | 65.74 ± 7.29 | 2.25 ± 0.43 |  |
| Weight (kg) | 1,540 | 83.14 ± 13.11 | 1,540 | 82.94 ± 13.14 | −0.20 ± 3.80 | 0.3075 |
| Height (cm) | 1,540 | 176.54 ± 6.65 |  |  |  |  |
| BMI (kg/m<sup>2</sup>) | 1,540 | 26.66 ± 3.82 | 1,540 | 26.60 ± 3.85 | −0.06 ± 1.21 | 0.3036 |
| Waist Circumference (cm) | 1,466 | 93.69 ± 10.71 | 1,537 | 94.01 ± 10.59 | 0.35 ± 5.89 | 0.0082 |
| Hip Circumference (cm) | 1,466 | 100.32 ± 7.35 | 1,537 | 99.69 ± 7.08 | −0.59 ± 5.22 | 0.00012* |
| Waist-to-Hip Ratio | 1,466 | 0.93 ± 0.06 | 1,537 | 0.94 ± 0.07 | 0.0091 ± 0.05 | 9.7 × 10<sup>−12</sup>* |
| Systolic BP (mm Hg) | 1,052 | 141.45 ± 16.19 | 1,398 | 144.09 ± 17.29 | 3.01 ± 14.00 | 6 × 10<sup>−11</sup>* |
| Diastolic BP (mm Hg) | 1,052 | 79.99 ± 9.61 | 1,398 | 81.35 ± 9.71 | 1.59 ± 8.34 | 4.4 × 10<sup>−09</sup>* |
| Grip Strength (kg) | 1,455 | 36.81 ± 8.38 | 1,533 | 34.75 ± 8.91 | −2.22 ± 7.29 | 3.6 × 10<sup>−28</sup>* |

The average duration between the first and second imaging visit was 2.25 ± 0.43 years for male participants and 2.27± 0.44 years for females (range 2.01 *−* 2.97 years across all participants). Despite some large individual changes in BMI between the first and second imaging visits (for visual examples, see Figure 1), there was no overall change in BMI, body weight or waist circumference (Table 3). The results showed small, though statistically significant, increases in waist-to-hip ratio (arising mainly from changes in hip circumference) and both systolic and diastolic blood pressure, along with a significant decrease in grip strength. These changes were mirrored in both male and female participants. While in absolute terms most of these changes are small and in some cases may not have physiological significance, the percentage decrease in grip strength, *−*2.3% in women and *−* 4.3% in men, in only two years is surprising and may have significant functional impact.

**Figure 1.**
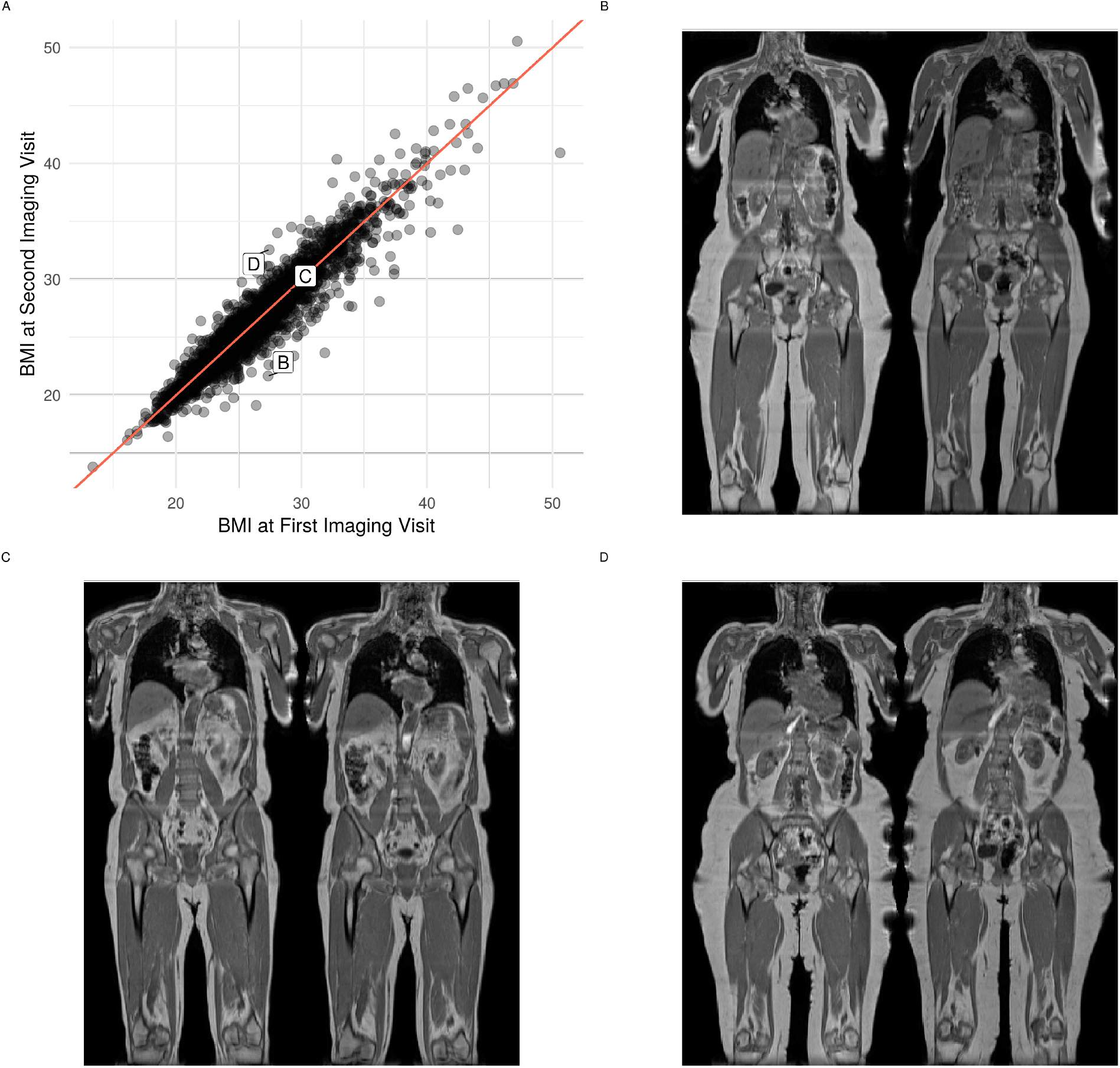
Change in BMI between the first and second imaging visits. (A) scatterplot of BMI values for all participants, (B) coronal view of participant whose BMI decreased by 5.7 kg/m^2^ from the first (left panel) to the second imaging visit (right panel), (C) coronal view of participant whose BMI did not change and (D) coronal view of participant whose BMI increased by 5.1 kg/m^2^.

In addition to the volumetric assessment of multiple abdominal organs (liver, pancreas, spleen and kidneys), changes in adipose tissue, skeletal muscle and the vascular system were also assessed. Furthermore, levels of PDFF and iron concentration of the liver, pancreas, paraspinal muscles and spleen were measured for each participant. Table 4 summarizes the IDPs at the first imaging visit for all participants and also separately by gender. We have previously reported gender differences in IDPs in the larger UK Biobank cohort^14^, and a similar pattern is observed in the longitudinal cohort. Most volume measurements (total skeletal muscle, kidney, liver, pancreas, heart, spleen and VAT) being larger in male participants, while ASAT volumes are larger in women. Organ iron levels were similar between genders, whereas organ PDFF was generally higher in men compared to women.

**Table 4.** Summary of IDPs at the first imaging visit for all participants, and separated by gender. Values are reported as mean and standard deviation. ASAT: abdominal subcutaneous adipsose tissue; Conc: concentration; CSA: cross-sectional area; PDFF: proton density fat fraction; VAT: visceral adipose tissue.

|  | *N* | All Participants | Women | Men |
| --- | --- | --- | --- | --- |
| Liver Volume (ml) | 2,919 | 1,400.61 ± 276.70 | 1,282.60 ± 217.11 | 1,519.01 ± 279.52 |
| Lungs Volume (ml) | 2,918 | 2,601.53 ± 714.94 | 2,238.47 ± 498.36 | 2,966.08 ± 714.09 |
| Pancreas Volume (ml) | 2,809 | 61.10 ± 15.27 | 57.07 ± 13.62 | 65.14 ± 15.77 |
| Left Kidney Volume (ml) | 2,916 | 145.30 ± 33.43 | 127.70 ± 25.56 | 163.00 ± 30.98 |
| Right Kidney Volume (ml) | 2,916 | 145.67 ± 30.88 | 130.42 ± 23.48 | 160.99 ± 29.83 |
| Spleen Volume (ml) | 2,913 | 170.22 ± 62.78 | 143.35 ± 45.26 | 197.18 ± 66.27 |
| ASAT Volume (ml) | 2,919 | 8,286.55 ± 3,923.02 | 9,669.30 ± 4,040.36 | 6,899.06 ± 3,258.31 |
| VAT Volume (ml) | 2,920 | 3,821.62 ± 2,226.94 | 2,688.03 ± 1,477.54 | 4,958.33 ± 2,272.45 |
| Total Thigh Subcutaneous Fat (ml) | 2,919 | 7,357.26 ± 2,970.84 | 8,895.89 ± 2,873.70 | 5,813.35 ± 2,153.87 |
| Total Thigh Internal Fat Volume (ml) | 2,919 | 655.60 ± 218.17 | 600.58 ± 176.14 | 710.80 ± 241.18 |
| Total Muscle Volume (ml) | 2,919 | 18,933.38 ± 4,473.99 | 15,134.07 ± 1,766.79 | 22,745.72 ± 2,818.74 |
| Total Iliopsoas Muscle Volume (ml) | 2,912 | 638.04 ± 170.82 | 498.53 ± 70.43 | 778.31 ± 119.51 |
| Total Paraspinal Muscle CSA (mm<sup>2</sup>) | 2,908 | 1,328.77 ± 491.79 | 1,066.77 ± 317.48 | 1,592.22 ± 495.32 |
| Total Thigh Muscle Volume (ml) | 2,919 | 8,647.08 ± 2,209.44 | 6,814.87 ± 964.52 | 10,485.58 ± 1,447.97 |
| Heart Volume (ml) | 2,918 | 498.74 ± 119.82 | 416.15 ± 66.77 | 581.66 ± 102.82 |
| Superior Vena Cava CSA (mm<sup>2</sup>) | 2,778 | 509.63 ± 118.01 | 472.22 ± 104.41 | 546.65 ± 119.05 |
| Descending Suprarenal Aorta CSA (mm<sup>2</sup>) | 2,888 | 643.58 ± 100.35 | 591.21 ± 81.52 | 696.10 ± 89.43 |
| Descending Thoracic Aorta CSA (mm<sup>2</sup>) | 2,891 | 643.36 ± 106.23 | 589.03 ± 84.41 | 697.80 ± 97.66 |
| Aortic Arch CSA (mm<sup>2</sup>) | 2,767 | 878.30 ± 199.51 | 842.40 ± 193.35 | 915.66 ± 199.03 |
| Descending Infrarenal Aorta CSA (mm<sup>2</sup>) | 2,871 | 390.22 ± 66.45 | 370.04 ± 58.36 | 410.38 ± 67.93 |
| Median Liver Iron Conc (mg/g) | 2,909 | 1.23 ± 0.31 | 1.21 ± 0.25 | 1.24 ± 0.36 |
| Median Pancreas Iron Conc (mg/g) | 2,689 | 0.77 ± 0.09 | 0.80 ± 0.10 | 0.75 ± 0.07 |
| Median Paraspinal Muscle Iron Conc (mg/g) | 2,908 | 1.18 ± 0.11 | 1.18 ± 0.13 | 1.18 ± 0.10 |
| Median Spleen Iron Conc (mg/g) | 2,753 | 0.90 ± 0.29 | 0.86 ± 0.27 | 0.94 ± 0.31 |
| Median Liver PDFF (%) | 2,909 | 4.72 ± 4.67 | 3.94 ± 4.05 | 5.50 ± 5.11 |
| Median Pancreas PDFF (%) | 2,689 | 10.10 ± 7.53 | 7.97 ± 5.90 | 12.22 ± 8.34 |
| Median Paraspinal Muscle PDFF (%) | 2,908 | 5.34 ± 2.29 | 5.69 ± 2.40 | 4.98 ± 2.12 |

Similar to the anthropometric parameters, in absolute terms, changes in the IDPs between the two visits were limited (Table 5) and included reduction in skeletal muscle, pancreas, liver and spleen volume (ranging *−* 1.3 to *−* 0.4%). No significant changes in liver and pancreas PDFF or iron concentration were observed. The largest and most consistent changes were observed in specific body-fat depots, with increases in VAT and intermuscular fat in the thighs (4.1% and 3.9%, respectively) taking place in the absence of any detectable changes in levels of subcutaneous fat in the abdomen or thighs.

**Table 5.**
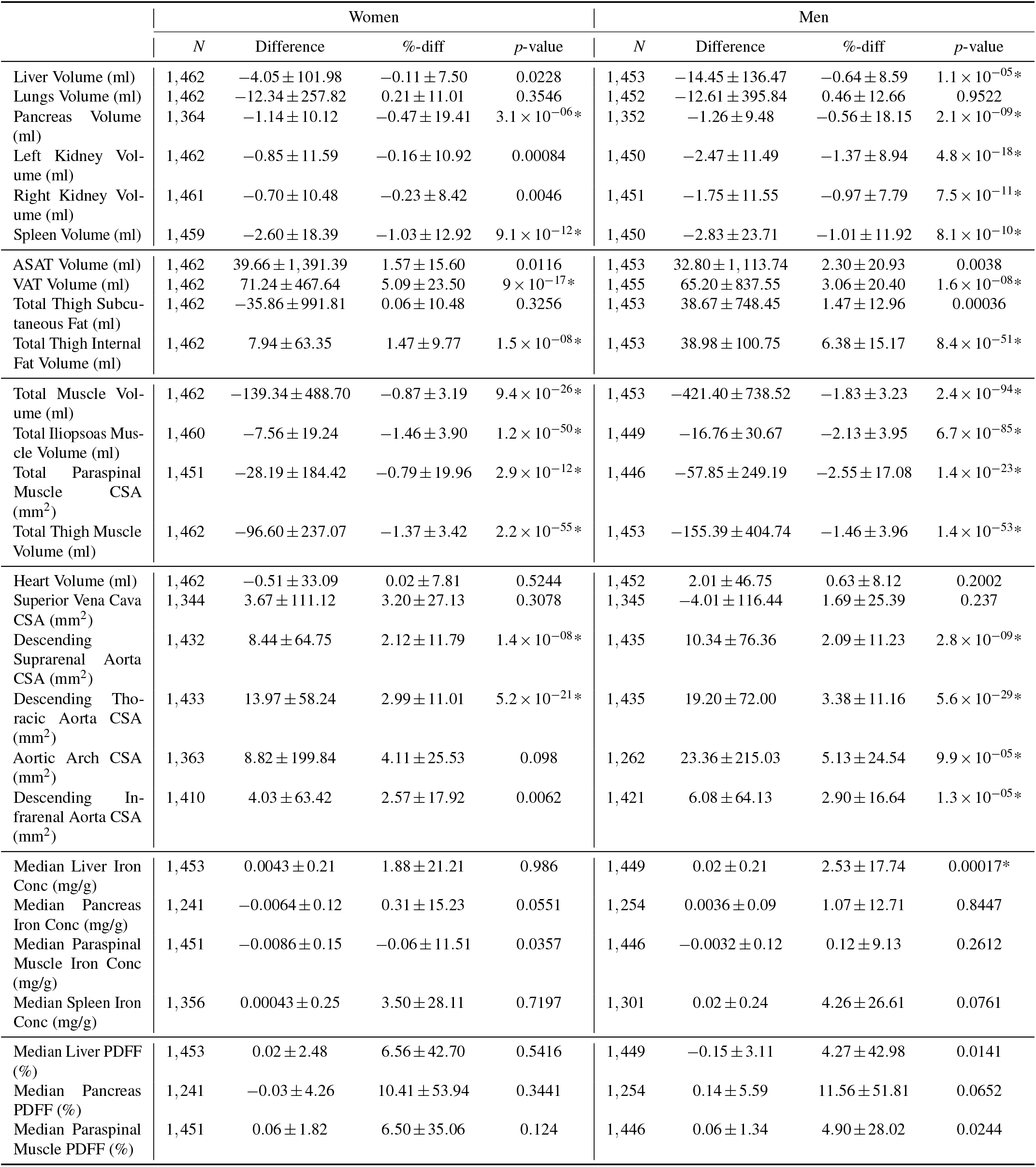
Change in IDPs between the first and second imaging visits. Values are reported as mean and standard deviation. An asterisk (*) indicates the *p*-value is below the significance threshold adjusted for multiple comparisons. ASAT: abdominal subcutaneous adipsose tissue; Conc: concentration; CSA: cross-sectional area; PDFF: proton density fat fraction; VAT: visceral adipose tissue.

To determine the potential impact of specific conditions on the longitudinal changes described above, the participants were categorised at their first imaging visit utilising standard ICD9/ICD10 and self-reported codes (Table 1 and Supplementary Table S2). A total of eight different conditions were identified, including type-1 and-2 diabetes, liver and kidney disease, CVD and cancer, with four of them being assessed against IDPs (Table 2). We adjusted for BMI, age, gender, body fat, blood pressure and grip strength, but found that chronic diseases did not result in statistically-significant increases or decreases at the second imaging visit in any of the IDPs, as measured by the interaction term between diagnosis at the first and second imaging visits (the last four rows of Figure 2 and Supplementary Table S3). However, we note that T2DM was associated with a reduction in total skeletal muscle (Model 1, *p* = 0.0130) and total iliopsoas muscle (Model 2, *p* = 0.0439) volumes at the second imaging visit, and a diagnosis of metabolic disorder was associated with a reduction in liver (Model 5, *p* = 0.0124) and total lung (Model 6, *p* = 0.0006) volume. Liver disease was linked to a reduction in total kidney volume (Model 7, *p* = 0.0219). We note that an increase in average liver volume of approximately 15 ml (Model 5) was observed for all subjects at the second imaging visit and achieved statistical significance (*p* = 3.93 *×* 10^*−*08^). The adjusted intraclass correlation coefficients for IDPs in the linear mixed-effects models (Supplementary Table S3) showed a high degree of repeatability for muscle and kidney volumes (0.92 to 0.96) and only slightly lower in the liver (0.84) and total lung (0.83) volumes.

**Figure 2.**
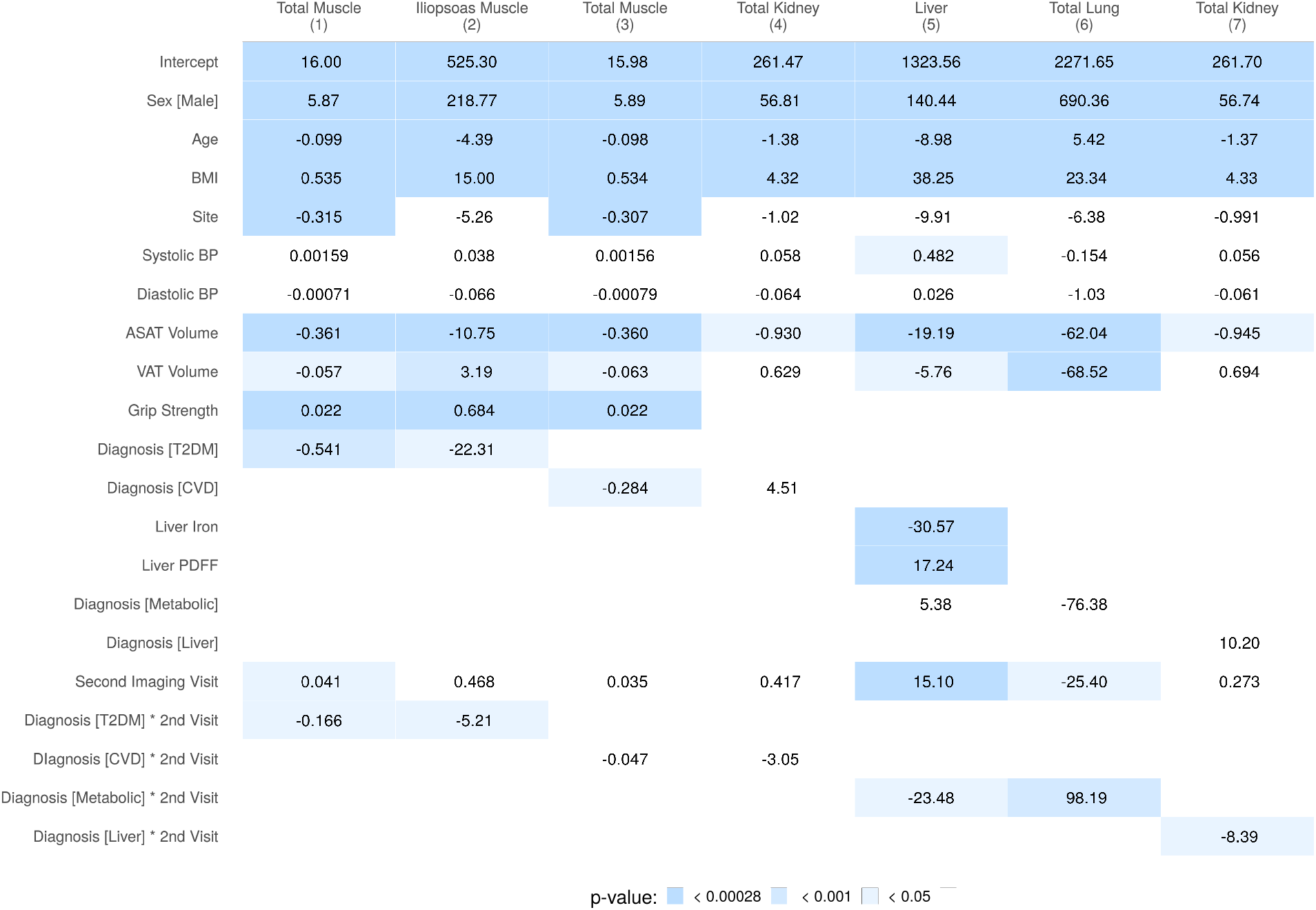
Summary of regression coefficients for the mixed-effects models in Table 2. All dependent variables are volumes measured in milliliters (ml), execept for total muscle (Models 1 and 3), ASAT and VAT which are in liters (l). ASAT: abdominal subcutaneous adipsose tissue; BMI: body mass index; BP: blood pressure; CVD: cardiovascular disease; PDFF: proton density fat fraction; T2DM: type-2 diabetes mellitus; VAT: visceral adipose tissue.

## Discussion

The UK Biobank is a unique resource originally designed to enable the in-depth study of a large cohort of the UK population in a cross-sectional manner^6,8^. The inclusion of a rescanning subprogram has presented an unparalleled opportunity to assess longitudinal changes in a free-living cohort. In this study we generated multiple organ IDPs^14^ and examined whether these changed over the study period (*<* 3 years). We also assessed if these changes were impacted upon by previously-diagnosed chronic conditions: cardiovascular, metabolic, liver and kidney diseases and cancer. While the interval between scans was relatively short, which does not allow delineation of changes directly arising from aging, this cohort can become an invaluable resource to understand short-term multimorbidity associated with free living in an obesogenic environment^26,27^.

Although we observed small differences in some anthropometric measurements and blood pressure across genders, these changes appear too small, in absolute terms, to be of great clinical significance^28^. However, larger changes in grip strength were observed across the whole cohort, in parallel to a decrease in total skeletal muscle volume and an increase in intramuscular fat infiltration. This suggests that even a relatively short period of time, under standard lifestyle conditions, can have significant repercussions on muscle quality and function and may be a precursor to subsequent health conditions^29^. A small decrease in hip circumference over the two-year period was an interesting observation. Previous cross-sectional studies have reported age-related increases in hip circumference up to 60 years of age, after which it decreases, whereas waist circumference continually increases with age^30^. This is clearly echoed in our study. The reduction in hip circumference in the elderly may reflect peripheral muscle wasting and given the age of many of the participants in our cohort rapid muscle loss is a distinct possibility. Indeed, the rate of age-related muscle loss is at its maximum beyond the age of 70 years^31^. The significant reductions in thigh muscle volume, also observed in our study, supports the possibility that changes in hip circumference relates to overall muscle wasting during this period of time.

In terms of changes in body composition arising from IDP measures, the cohort as a whole showed a significant increase in visceral fat content, accompanied by an increase in intramuscular fat content. This despite no changes in BMI, waist circumference and abdominal subcutaneous adipose tissue depot. The unadjusted changes in VAT were also accompanied by reductions in all skeletal muscle IDPs including total, iliopsoas and thigh muscle volumes. Again while relatively small in absolute terms (*−* 1.8 to *−* 1.3%), these changes were consistent across genders. An increase in internal fat depots, accompanied by a reduction in muscle volume, are usually observed in aging cohorts and are generally connected to a deleterious body composition, which in turn is associated with poorer health outcomes^32–35^. However, the changes observed in this study took place over a relatively short period of time, probably reflecting the combination of decreased activity common in this age group coupled with the ongoing impact of the obesogenic environment inhabited by the participants^36,37^, rather than ageing itself^38,39^. In parallel to the changes in internal fat, we observed small but significant reductions in pancreas volume in both genders. Although large-scale studies of pancreatic volume are relatively limited, there is consistent evidence suggesting that both type-1 and -2 diabetes are associated with reduced pancreas volume^40–42^, with some studies suggesting that remission of type-2 diabetes through calorie restriction results in an increase in pancreatic volume^43^. Thus, while the reduction in pancreas volume is small, it does seem to fit with the overall picture of a less healthy body composition. We also observed small reductions in kidney volume, although these only reached statistical significance in men. Decreases in kidney volume have been reported as a function of age, reduced kidney volume is associated with indices of poorer renal function^44,45^. Similarly, we observed small yet significant increases in the cross-sectional area of the descending aorta (suprarenal, thoracic) in both men and women, with additional increases in infrarenal descending aorta and aortic arch in men. Increased size of coronary arteries, and in particular the descending aorta, has been previously reported as an adaptive response to ensure adequate blood flow in aortic valve conditions^46,47^. This observation adds further weight to the argument that collectively, the overall observed changes in IDPs in this cohort reflect worsening phenotypes over a relatively short period of time. However, after adjusting for potential confounding factors, many of the observed changes were no longer apparent in the wider cohort. We sought to question whether the presence of chronic disease such as diabetes, metabolic disorders, CVD, liver and kidney disease as well as cancer, would accelerate or indeed ameliorate changes in IDPs over the two year follow-up period. We hypothesised that the changes to organs and tissues would be amplified in the presence of preexisting conditions, however, this was not the case. Given the relatively small numbers of individuals diagnosed with the diseases of interest, the relatively short scan interval and without additional information regarding where in the disease process they were/mitigating effects of treatment etc it is not possible to conclusively determine whether chronic diseases have a discernible impact on organ volume/composition.

Previous cross-sectional and longitudinal studies have shown a strong correlation between VAT and ectopic fat depots^33^, suggesting changes in the former should have been reflected in the latter, especially liver fat. However, the expected changes in ectopic fat were not observed in the current study. This discordance may reflect yet unidentified underlying mechanisms associated with the deposition of fat in different depots^48^ or reflect the heterogeneity and variability in fat deposition in ectopic depots and thus be more susceptible to the size of the cohort^49^. Indeed, though statistical changes in PDFF did not reach the Bonferroni corrected significance threshold, we did observe consistent increases in both the liver (6.6% in women and 4.3% in men) and pancreas (10.4% in women and 11.6% in men). Whilst care must be taken in the interpretation of the above observations, they do fit with the overall pattern of changes we observed elsewhere in the body. Completion of the re-scanning pilot (*N* = 10, 000) should substantially help to resolve some of these inconsistencies.

The current study has a number of limitations. First, the overall cohort is relatively healthy compared to the general population and thus may not be fully representative of the UK population, nor does it include data from younger participants (*<* 40 years). A second limitation of the current study relates to the possible clinical impact of some of the changes we have observed, while statistically significant and of scientific interest they are relatively small in magnitude. We would expect that in a cohort with a greater incidence of chronic disease the changes reported here would be substantially larger. This point may be addressed in the future when the whole UK Biobank imaging cohort is rescanned, with a longer time lapse between scans (*>* 6 years), or by scanning other biobank populations enriched with a larger number of participants with relevant clinical conditions. Similarly, whilst we used accurate IDPs, no biochemical data was available to improve our understanding of the clinical and metabolic significance of the findings. Furthermore, the reported IDPs represent organs as a whole, ignoring the possibility of detecting within-organ heterogeneity. Finally, though no reproducibility measurements are currently available in the UK Biobank, our results arise from group averages instead of measurements on a single subject. All the changes are in the expected direction for an ageing population, giving confidence to their statistical significance. Future studies with a larger population, scanned over a longer interval with detailed metabolic profiles could help yield more precise results which will allow a greater understanding regarding the clinical implications of the changes observed here.

In conclusion, the use of image derived phenotypes allowed us to detect some significant longitudinal changes in body composition after a relatively short period of time. The expected increase in sample size should help to confirm some of these preliminary results.

## Data Availability

Researchers may apply to use the UKBB data resource by submitting a health-related research proposal that is in the public interest. More information may be found on the UKBB researchers and resource catalogue pages (www.ukbiobank.ac.uk).

## Acknowledgements

This study was carried out using UK Biobank Application number 44584, and we thank the participants in the UK Biobank imaging study. This study was funded by Calico Life Sciences LLC. We thank Dr. Amoolya Singh and Dr. Kevin Wright for comments that improved the manuscript.

## Author contributions

JDB, ELT, BW, MC and YL conceived the study. JDB, BW, ELT, NB and MT designed the study. NB, BW, EPS, MC and YL implemented the methods and performed the data analysis. MT defined the disease and physiological condition categories. BW performed the statistical analysis. ELT, BW, MT and NB drafted the manuscript. All authors read and approved the manuscript.

## Additional information

**Supplementary information** is available online.

## References

1. Mahmood, S. S., Levy, D., Vasan, R. S. & Wang, T. J. The Framingham Heart Study and the epidemiology of cardiovascular disease: a historical perspective. The Lancet 383, 999–1008, DOI: 10.1016/s0140-6736(13)61752-3 (2014).

2. Lee, D. H. & Giovannucci, E. L. Body composition and mortality in the general population: A review of epidemiologic studies. Exp. Biol. Medicine 243, 1275–1285, DOI: 10.1177/1535370218818161 (2018).

3. Rospleszcz, S. et al. Association of longitudinal risk profile trajectory clusters with adipose tissue depots measured by magnetic resonance imaging. Sci. Reports 9, DOI: 10.1038/s41598-019-53546-y (2019).

4. Volzke, H. et al. Cohort profile: The study of health in Pomerania. Int. J. Epidemiol. 40, 294–307, DOI: 10.1093/ije/dyp394 (2010).

5. Victor, R. G. et al. The Dallas Heart Study: a population-based probability sample for the multidisciplinary study of ethnic differences in cardiovascular health. The Am. J. Cardiol. 93, 1473–1480, DOI: 10.1016/j.amjcard.2004.02.058 (2004).

6. Sudlow, C. et al. UK Biobank: An open access resource for identifying the causes of a wide range of complex diseases of middle and old age. PLOS Medicine 12, e1001779, DOI: 10.1371/journal.pmed.1001779 (2015).

7. Bamberg, F. et al. Whole-body MR imaging in the German National Cohort: Rationale, design, and technical background. Radiology 277, 206–220, DOI: 10.1148/radiol.2015142272 (2015).

8. Littlejohns, T. J. et al. The UK Biobank imaging enhancement of 100,000 participants: rationale, data collection, management and future directions. Nat. Commun. 11, DOI: 10.1038/s41467-020-15948-9 (2020).

9. Lees, J. S. et al. Glomerular filtration rate by differing measures, albuminuria and prediction of cardiovascular disease, mortality and end-stage kidney disease. Nat. Medicine 25, 1753–1760, DOI: 10.1038/s41591-019-0627-8 (2019).

10. Tavaglione, F. et al. Inborn and acquired risk factors for severe liver disease in europeans with type 2 diabetes from the UK Biobank. JHEP Reports 3, 100262, DOI: 10.1016/j.jhepr.2021.100262 (2021).

11. Schneider, C. V., Zandvakili, I., Thaiss, C. A. & Schneider, K. M. Physical activity is associated with reduced risk of liver disease in the prospective UK Biobank cohort. JHEP Reports 3, 100263, DOI: 10.1016/j.jhepr.2021.100263 (2021).

12. Lin, B. M. et al. Genetics of chronic kidney disease stages across ancestries: The PAGE study. Front. Genet. 10, DOI: 10.3389/fgene.2019.00494 (2019).

13. Basty, N. et al. Automated measurement of pancreatic fat and iron concentration using multi-echo and T1-weighted MRI data. In 2020 IEEE 17th International Symposium on Biomedical Imaging (ISBI), DOI: 10.1109/isbi45749.2020.9098650 (2020).

14. Liu, Y. et al. Genetic architecture of 11 organ traits derived from abdominal MRI using deep learning. eLife 10, DOI: 10.7554/elife.65554 (2021).

15. Bydder, M. et al. Constraints in estimating the proton density fat fraction. Magn. Reson. Imaging 66, 1–8, DOI: 10.1016/j.mri.2019.11.009 (2020).

16. McKay, A. et al. Measurement of liver iron by magnetic resonance imaging in the UK Biobank population. PLOS ONE 13, e0209340, DOI: 10.1371/journal.pone.0209340 (2018).

17. Authors/Task Force Members et al. 2014 ESC Guidelines on the diagnosis and treatment of aortic diseases: Document covering acute and chronic aortic diseases of the thoracic and abdominal aorta of the adult the task force for the diagnosis and treatment of aortic diseases of the european society of cardiology (ESC). Eur. Hear. J. 35, 2873–2926, DOI: 10.1093/eurheartj/ehu281 (2014).

18. Grélard, F., Baldacci, F., Vialard, A. & Domenger, J.-P. New methods for the geometrical analysis of tubular organs. Med. Image Analysis 42, 89–101, DOI: 10.1016/j.media.2017.07.008 (2017).

19. R Core Team. R: A Language and Environment for Statistical Computing. R Foundation for Statistical Computing, Vienna, Austria (2020).

20. Bates, D., Mächler, M., Bolker, B. & Walker, S. Fitting linear mixed-effects models using lme4. J. Stat. Softw. 67, DOI: 10.18637/jss.v067.i01 (2015).

21. Harrison, L., Dunn, D. T., Green, H. & Copas, A. J. Modelling the association between patient characteristics and the change over time in a disease measure using observational cohort data. Stat. Medicine 28, 3260–3275, DOI: 10.1002/sim.3725 (2009).

22. Satterthwaite, F. E. An approximate distribution of estimates of variance components. Biom. Bull. 2, 110, DOI: 10.2307/3002019 (1946).

23. Kuznetsova, A., Brockhoff, P. B. & Christensen, R. H. B. lmerTest package: Tests in linear mixed effects models. J. Stat. Softw. 82, DOI: 10.18637/jss.v082.i13 (2017).

24. Nakagawa, S., Johnson, P. C. D. & Schielzeth, H. The coefficient of determination R<sup>2</sup> and intra-class correlation coefficient from generalized linear mixed-effects models revisited and expanded. J. The Royal Soc. Interface 14, 20170213, DOI: 10.1098/rsif.2017.0213 (2017).

25. Lüdecke, D., Ben-Shachar, M., Patil, I., Waggoner, P. & Makowski, D. performance: An R package for assessment, comparison and testing of statistical models. J. Open Source Softw. 6, 3139, DOI: 10.21105/joss.03139 (2021).

26. Chudasama, Y. V. et al. Physical activity, multimorbidity, and life expectancy: a UK Biobank longitudinal study. BMC Medicine 17, DOI: 10.1186/s12916-019-1339-0 (2019).

27. Chudasama, Y. V. et al. Healthy lifestyle and life expectancy in people with multimorbidity in the UK Biobank: A longitudinal cohort study. PLOS Medicine 17, e1003332, DOI: 10.1371/journal.pmed.1003332 (2020).

28. Pinto, E. Blood pressure and ageing. Postgrad. Med. J. 83, 109–114, DOI: 10.1136/pgmj.2006.048371 (2007).

29. Reeve, T. E. et al. Grip strength measurement for frailty assessment in patients with vascular disease and associations with comorbidity, cardiac risk, and sarcopenia. J. Vasc. Surg. 67, 1512–1520, DOI: 10.1016/j.jvs.2017.08.078 (2018).

30. Teh, B. H., Pan, W. H. & J, C. C. The reallocation of body fat toward the abdomen persists to very old age, while body mass index declines after middle age in Chinese. Int. J. Obes. Relat. Metab. Disord. 20, 683–687 (1996).

31. Mitchell, W. K. et al. Sarcopenia, dynapenia, and the impact of advancing age on human skeletal muscle size and strength: a quantitative review. Front. Physiol. 3, DOI: 10.3389/fphys.2012.00260 (2012).

32. Lauretani, F. et al. Age-associated changes in skeletal muscles and their effect on mobility: an operational diagnosis of sarcopenia. J. Appl. Physiol. 95, 1851–1860, DOI: 10.1152/japplphysiol.00246.2003 (2003).

33. Thomas, E. L. et al. The missing risk: MRI and MRS phenotyping of abdominal adiposity and ectopic fat. Obesity 20, 76–87, DOI: 10.1038/oby.2011.142 (2012).

34. Therkelsen, K. E. et al. Intramuscular fat and associations with metabolic risk factors in the Framingham Heart Study. Arter. Thromb. Vasc. Biol. 33, 863–870, DOI: 10.1161/atvbaha.112.301009 (2013).

35. Kalyani, R. R., Corriere, M. & Ferrucci, L. Age-related and disease-related muscle loss: the effect of diabetes, obesity, and other diseases. The Lancet Diabetes & Endocrinol. 2, 819–829, DOI: 10.1016/s2213-8587(14)70034-8 (2014).

36. Abarca-Gómez, L. et al. Worldwide trends in body-mass index, underweight, overweight, and obesity from 1975 to 2016: a pooled analysis of 2416 population-based measurement studies in 128·9 million children, adolescents, and adults. The Lancet 390, 2627–2642, DOI: 10.1016/s0140-6736(17)32129-3 (2017).

37. Swinburn, B. A. et al. The global obesity pandemic: shaped by global drivers and local environments. The Lancet 378, 804–814, DOI: 10.1016/s0140-6736(11)60813-1 (2011).

38. Owen, N., Sparling, P. B., Healy, G. N., Dunstan, D. W. & Matthews, C. E. Sedentary behavior: Emerging evidence for a new health risk. Mayo Clin. Proc. 85, 1138–1141, DOI: 10.4065/mcp.2010.0444 (2010).

39. Adams, J., Mytton, O., White, M. & Monsivais, P. Why are some population interventions for diet and obesity more equitable and effective than others? the role of individual agency. PLOS Medicine 13, e1001990, DOI: 10.1371/journal.pmed.1001990 (2016).

40. DeSouza, S. V. et al. Pancreas volume in health and disease: a systematic review and meta-analysis. Expert. Rev. Gastroenterol. & Hepatol. 12, 757–766, DOI: 10.1080/17474124.2018.1496015 (2018).

41. Saisho, Y. et al. Pancreas volumes in humans from birth to age one hundred taking into account sex, obesity, and presence of type-2 diabetes. Clin. Anat. 20, 933–942, DOI: 10.1002/ca.20543 (2007).

42. Lim, S. et al. Differences in pancreatic volume, fat content, and fat density measured by multidetector-row computed tomography according to the duration of diabetes. Acta Diabetol. 51, 739–748, DOI: 10.1007/s00592-014-0581-3 (2014).

43. Al-Mrabeh, A. et al. 2-year remission of type 2 diabetes and pancreas morphology: a post-hoc analysis of the DiRECT open-label, cluster-randomised trial. The Lancet Diabetes & Endocrinol. 8, 939–948, DOI: 10.1016/s2213-8587(20)30303-x (2020).

44. Roseman, D. A. et al. Clinical associations of total kidney volume: the Framingham Heart Study. Nephrol. Dial. Transplantation gfw237, DOI: 10.1093/ndt/gfw237 (2016).

45. Hricak, H. et al. Renal parenchymal disease: sonographic-histologic correlation. Radiology 144, 141–147, DOI: 10.1148/radiology.144.1.7089245 (1982).

46. O’keefe, J. H., Owen, R. M. & Bove, A. A. Influence of left ventricular mass on coronary artery cross-sectional area. The Am. J. Cardiol. 59, 1395–1397, DOI: 10.1016/0002-9149(87)90927-1 (1987).

47. Villari, B. et al. Regression of coronary artery dimensions after successful aortic valve replacement. Circulation 85, 972–978, DOI: 10.1161/01.cir.85.3.972 (1992).

48. Mantovani, A., Byrne, C. D., Bonora, E. & Targher, G. Nonalcoholic fatty liver disease and risk of incident type 2 diabetes: A meta-analysis. Diabetes Care 41, 372–382, DOI: 10.2337/dc17-1902 (2018).

49. Lonardo, A., Leoni, S., Alswat, K. A. & Fouad, Y. History of nonalcoholic fatty liver disease. Int. J. Mol. Sci. 21, 5888, DOI: 10.3390/ijms21165888 (2020).

