## Supplemental File for "Precision MRI Phenotyping Enables Detection of Small Changes in Body Composition for Longitudinal Cohorts"

### ABSTRACT

### Supplementary Information

| | Women | | Men | |
|---|---|---|---|---|
| | First Imaging Visit | $p$-value | First Imaging Visit | $p$-value |
| $N$ | 23,742 | | 22,167 | |
| $N$ (Cheadle) | 13,912 | | 13,399 | |
| $N$ (Newcastle) | 5,966 | | 5,311 | |
| Age | $63.20 \pm 7.52$ | $1.1 \times 10^{-05}*$ | $64.70 \pm 7.78$ | $2.4 \times 10^{-10}*$ |
| Weight (kg) | $69.49 \pm 13.48$ | 0.0572 | $84.12 \pm 13.77$ | 0.0132 |
| Height (cm) | $162.55 \pm 6.25$ | 0.2174 | $175.86 \pm 6.61$ | $8.5 \times 10^{-05}*$ |
| BMI (kg/m$^2$) | $26.30 \pm 4.89$ | 0.0025 | $27.18 \pm 4.03$ | $6.7 \times 10^{-07}*$ |
| Waist Circumference (cm) | $83.48 \pm 11.98$ | 0.0214 | $94.90 \pm 10.89$ | $9 \times 10^{-05}*$ |
| Hip Circumference (cm) | $101.54 \pm 9.96$ | 0.0349 | $101.26 \pm 7.38$ | $3.2 \times 10^{-06}*$ |
| Waist-to-Hip Ratio | $0.82 \pm 0.07$ | 0.4512 | $0.94 \pm 0.06$ | 0.2387 |
| Systolic BP (mm Hg) | $136.57 \pm 19.33$ | 0.0865 | $142.46 \pm 17.34$ | 0.1413 |
| Diastolic BP (mm Hg) | $77.59 \pm 9.91$ | $5.4 \times 10^{-08}*$ | $80.92 \pm 9.75$ | 0.0023 |
| Grip Strength (kg) | $21.50 \pm 5.78$ | 0.0535 | $36.17 \pm 8.58$ | 0.00068 |

**Table S1.** All participants with an initial imaging visit for the Cheadle and Newcastle sites. P-values represent group comparisons with the longitudinal cohort from Table 3. An asterisk (*) indicates the $p$-value is below the significance threshold adjusted for multiple comparisons. BMI: body mass index; BP: blood pressure.

| Trait | SNOMED | ICD9 | ICD10 | Field 20002 | Field 20001 | Other Fields |
|---|---|---|---|---|---|---|
| Cardiovascular disease (defined by myocardial infraction, stroke and heart failure) | 22298006 230690007 84114007 | 410-412\|4289 | I20-I25\|I60-I64 | 1074\|1075\|1076\|1081\|1086\|1491\|1583 | | |
| Liver disease (defined by chronic viral hepatitis, malignant liver disease, NAFLD and hepatic cirrhosis) | 235856003 3738000 197315008 19943007 | 0700\|0702\|0703\|0704\|0709\|155\|1551\|5715\|57151\|5716\|5718\|573\|5731\|5734 | B18\|C22\|K70-K77 | 1136\|1156\|1158\|1604 | | |
| Kidney disease (defined by chronic kidney disease and end-stage renal disease) | 709044004 46177005 | 585\|5859 | N180-N185\|N188\|N189 | 1192\|1193 | | |
| Type 1 diabetes mellitus | 46635009 | 25001 | E10\|E100-E109 | 1222 | | |
| Type 2 diabetes mellitus | 44054006 | 25000 | E11\|E110-E119 | 1220\|1223 | | |
| Cancer (all types) | 363346000 | Type of cancer ICD9 (Field 40013) | Type of cancer ICD10 (Field 40006) | | All cancer codes | 2453 |
| Metabolic disorder (correspond to carbohydrates and lipids) | 75934005 | 272 | E70-E90 | | | |
| Menopause | 289903006 | 6272 | N951 | 1665 | | 2724 |

**Table S2.** Definitions for disease/physiological conditions for cardiovascular disease, liver disease, kidney disease, type-1 and type-2 diabetes mellitus, any cancer, metabolic disorder and menopause. Field 20001: self-reported cancer illness; Field 20002: self-reported non-cancer illness; ICD9: International Classification of Diseases 9th edition; ICD10 International Classification of Diseases 10th edition.

| | *Dependent variable:* | | | | | | |
|---|---|---|---|---|---|---|---|
| | Total Muscle | Iliopsoas Muscle | Total Muscle | Total Kidney | Liver | Total Lung | Total Kidney |
| | (1) | (2) | (3) | (4) | (5) | (6) | (7) |
| Intercept | 16.00*** | 525.30*** | 15.98*** | 261.47*** | 1,323.56*** | 2,271.65*** | 261.70*** |
| | (0.07) | (3.19) | (0.07) | (1.75) | (7.48) | (22.20) | (1.74) |
| Sex [Male] | 5.87*** | 218.77*** | 5.89*** | 56.81*** | 140.44*** | 690.36*** | 56.74*** |
| | (0.10) | (4.63) | (0.10) | (2.60) | (11.17) | (33.16) | (2.60) |
| Age | −0.10*** | −4.39*** | −0.10*** | −1.38*** | −8.98*** | 5.42*** | −1.37*** |
| | (0.005) | (0.22) | (0.005) | (0.12) | (0.49) | (1.46) | (0.12) |
| BMI | 0.53*** | 15.00*** | 0.53*** | 4.32*** | 38.25*** | 23.34*** | 4.33*** |
| | (0.02) | (0.65) | (0.02) | (0.39) | (1.73) | (5.13) | (0.39) |
| Site | −0.32*** | −5.26 | −0.31*** | −1.02 | −9.91 | −6.38 | −0.99 |
| | (0.07) | (3.17) | (0.07) | (1.67) | (6.80) | (20.19) | (1.67) |
| Systolic BP | 0.002 | 0.04 | 0.002 | 0.06 | 0.48* | −0.15 | 0.06 |
| | (0.001) | (0.05) | (0.001) | (0.03) | (0.17) | (0.53) | (0.03) |
| Diastolic BP | −0.001 | −0.07 | −0.001 | −0.06 | 0.03 | −1.03 | −0.06 |
| | (0.002) | (0.08) | (0.002) | (0.06) | (0.31) | (0.94) | (0.06) |
| ASAT Volume | −0.36*** | −10.75*** | −0.36*** | −0.93* | −19.19*** | −62.04*** | −0.94* |
| | (0.02) | (0.66) | (0.02) | (0.41) | (1.84) | (5.50) | (0.41) |
| VAT Volume | −0.06* | 3.19** | −0.06* | 0.63 | −5.76* | −68.52*** | 0.69 |
| | (0.02) | (0.96) | (0.02) | (0.58) | (2.62) | (7.52) | (0.58) |
| Grip Strength | 0.02*** | 0.68*** | 0.02*** | | | | |
| | (0.002) | (0.08) | (0.002) | | | | |
| Diagnosis [T2DM] | −0.54** | −22.31* | | | | | |
| | (0.16) | (7.55) | | | | | |
| Diagnosis [CVD] | | | −0.28* | 4.51 | | | |
| | | | (0.14) | (3.43) | | | |
| Liver Iron | | | | | −30.57*** | | |
| | | | | | (7.99) | | |
| Liver PDFF | | | | | 17.24*** | | |
| | | | | | (0.67) | | |
| Diagnosis [Metabolic] | | | | | 5.38 | −76.38 | |
| | | | | | (14.90) | (44.41) | |
| Diagnosis [Liver] | | | | | | | 10.20 |
| | | | | | | | (7.71) |
| Second Imaging Visit | 0.04* | 0.47 | 0.03 | 0.42 | 15.10*** | −25.40* | 0.27 |
| | (0.02) | (0.75) | (0.02) | (0.52) | (2.74) | (8.30) | (0.51) |
| Diagnosis [T2DM] * 2nd Visit | −0.17* | −5.21* | | | | | |
| | (0.07) | (2.59) | | | | | |
| DIagnosis [CVD] * 2nd Visit | | | −0.05 | −3.05 | | | |
| | | | (0.05) | (1.65) | | | |
| Diagnosis [Metabolic] * 2nd Visit | | | | | −23.48* | 98.19** | |
| | | | | | (9.38) | (28.59) | |
| Diagnosis [Liver] * 2nd Visit | | | | | | | −8.39* |
| | | | | | | | (3.66) |
| Participants | 2,974 | 2,966 | 2,974 | 2,989 | 2,993 | 2,994 | 2,989 |
| Adjusted ICC | 0.95 | 0.96 | 0.95 | 0.92 | 0.84 | 0.83 | 0.92 |
| BIC | 15,141 | 50,247 | 15,159 | 47,126 | 61,762 | 72,446 | 47,121 |

*Note:* *p<0.05; **p<0.001; ***p<0.00028

**Table S3.** Summary of regression coefficients for the linear mixed-effects models in Table 2. Standard errors are provided underneath the regression coefficients. Indicators are appended to the regression coefficients when the *p*-value is less than 0.05 *, 0.001 ** and 0.00028 *** (Bonferroni corrected). All volumes are in milliliters (ml), execept for total muscle, ASAT and VAT which are in liters (l). ASAT: abdominal subcutaneous adipsose tissue; BIC: Bayesian information criterion; BMI: body mass index; BP: blood pressure; CVD: cardiovascular disease; ICC: intraclass correlation coefficient; PDFF: proton density fat fraction; T2DM: type-2 diabetes mellitus; VAT: visceral adipose tissue.
